# Utilizing Saliva for Non-Invasive Detection of Exercise-Induced Myocardial Injury with Point-of-Care Cardiac Troponin-I

**DOI:** 10.1101/2024.12.02.24318327

**Authors:** Aleksandr N. Ovchinnikov

## Abstract

**Background:** Point-of-care (POC) cardiac troponin-I (cTnI) measurement methods often involve immunoassays, which can provide a momentary view of cTnI levels but the current modality highly restricts access to and frequency of testing in a sports and exercise medicine setting due to the requirement of a blood draw.

**Objectives:** This study aimed to compare cTnI concentrations in saliva and serum in athletes before (T1), early (T2), 4 h (T3), and 24 h (T4) after exercise.

**Methods:** 82 male runners were recruited and then divided into two groups. 54 participants (group 1) completed a 5-km time-trial, while 28 participants (group 2) did not undergo this exercise. POC testing device was used to quantify salivary and serum concentrations of cTnI in both groups at T1, T2, T3, and T4.

**Results:** In group 1, salivary and serum concentrations of cTnI increased at T2 (0.41±0.06 ng/mL and 0.48±0.06 ng/mL) compared to T1 (0.18±0.04 ng/mL and 0.22±0.04 ng/mL), reaching the highest values at T3 (0.62±0.05 ng/mL and 0.76±0.05 ng/mL) with the subsequent return to baseline values at T4 (0.16±0.03 ng/mL and 0.22±0.03 ng/mL). In group 2, there were no time-dependent changes in cTnI levels in both saliva (T1: 0.17±0.04 ng/mL, T2: 0.16±0.03 ng/mL, T3: 0.16±0.04 ng/mL, T4: 0.16±0.04 ng/mL) and serum (T1: 0.22±0.04 ng/mL, T2: 0.22±0.04 ng/mL, T3: 0.21±0.03 ng/mL, T4: 0.21±0.04 ng/mL). Salivary and serum concentrations of cTnI were significantly lower in group 2 compared to group 1 at T2 and T3; there was no difference between groups at T1 and T4. Deming regression and Passing–Bablok regression revealed that there was differential bias (at T3), but proportional agreement (at T1, T2, T3, T4) between salivary and serum levels of cTnI in both groups. The Bland–Altman method indicated that there was a negative differential bias but no proportional bias in the data. Recalibration of the new measurement approach (measurement of cTnI levels in saliva) by using the MethodCompare R package was effective in removing existing bias, as evidenced by its similar precision to the reference method (measurement of cTnI levels in serum), particularly at T2, T3, and T4.

**Conclusions:** In athletic settings, quantification of cTnI levels in saliva utilizing the POC-cTnI-Getein1100 assay may be a useful non-invasive tool in evaluating whether exercise-induced increases in cTnI levels are transient or there are acutely or chronically elevated cTnI concentrations.

**Graphical abstract:** 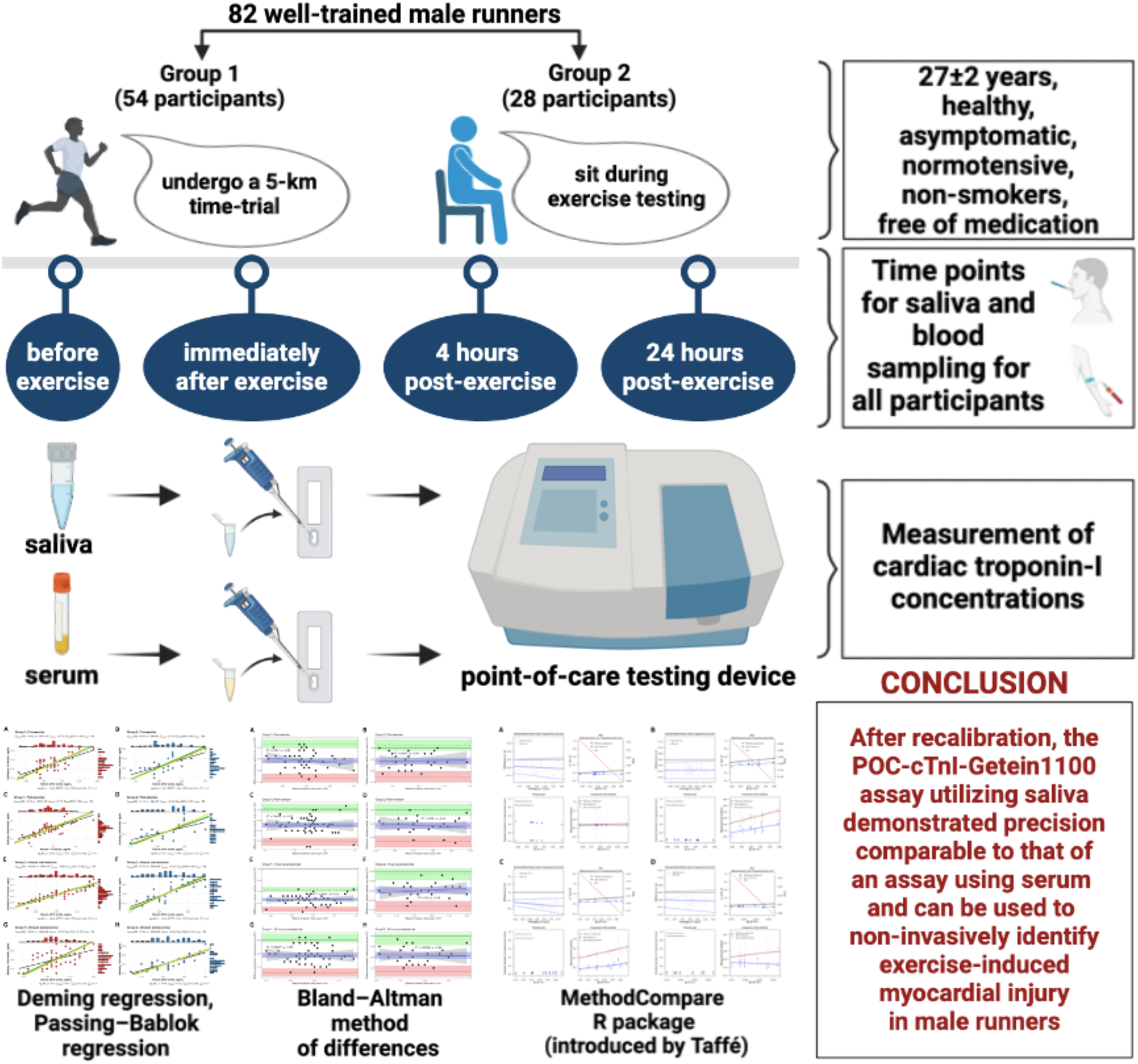

## Introduction

Over 60% of all fatalities during sports are cardiovascular related [1–3]. Sudden cardiac death (SCD) is the leading cause of mortality in athletes, including young athletes with “no health issues”, during exercise or in its aftermath [1–6]. Furthermore, over 70% of affected individuals are asymptomatic (or mildly symptomatic) before SCD [2, 7, 8]. Estimates of the rate of SCD in athletes can differ greatly (>300-fold) depending on the sport and population studied [6]. A portion of SCD cases is attributed to acute myocardial infarction (AMI), which is defined by identification of the rise and/or drop in cardiac markers (preferably cardiac troponin-I) with at least 1 value above the 99th percentile of the upper reference limit, along with evidence of myocardial ischemia (based on electrocardiography and/or magnetic resonance imaging) [9, 10, 11]. In spite of tremendous progress in the development of new diagnostic and screening methods, many AMI cases are still missed or diagnosed too late to be treated effectively [10, 12, 13]. Consequently, early detection of any unnecessary myocardial injury and its follow-up assessment in exercising individuals, including elite athletes, is essential to prevent severe outcomes and ensure long-term cardiac health.

Due to the potential for analysis outside the laboratory, it is becoming increasingly attractive to use quantitative point-of-care (POC) assays, which eliminate the need to deliver samples to the lab and, thereby, shorten assay times [14, 15]. However, despite the documented reduction in turnaround time for cardiac marker results through the use of POC approaches, just a fraction of emergency rooms worldwide has implemented these approaches [13, 16]. A perceived lack of precision, concerns about standardization, and difficulties with clinical data management are frequently cited reasons for the slow adoption rate of these new promising POC cardiac diagnostic options [13]. Because of these reasons and recent improvements in ultrasensitive lab-based assays, cardiac troponin (cTn) is considered as the preferred marker in serum in relation to myoglobin and creatine kinase-MB [13, 17].

Since the 1990s, cTn testing has enabled prompt and accurate assessment of cardiac injury, making it a useful tool in both clinical and sports settings [18–23]. The presence of detectable cTn in the blood following moderate to strenuous exercise in individuals without heart disease is well described in the literature [20–27]. Similar findings have been replicated across athletes representing a wide variety of endurance and team-sport discipline [22, 23, 28, 29]. Although athletes may experience transient cTn elevations depending on the intensity and duration of exercise, not all elevated states of cTn are created equal [7, 22, 23]. Elevated cTn levels may be associated with an increased risk of adverse cardiac events in exercising individuals specifically when prolonged (>24 h) elevation above a specific cut-off point (i.e., the 99th percentile value) is observed after exercise [7, 10, 22, 26]. Moreover, the risk of SCD may correlate with the severity of myocardial inflammation and serum concentrations of cTn [7, 30]. Therefore, as with any diagnostic tool, the pitfalls of clinical application deserve consideration.

As of now, cTn testing, when coupled with the appropriate clinical scenario, is the gold-standard method of documenting acute myocardial injury [19–29, 31–33]. POC troponin measurement methods often involve immunoassays based on the use of two or more antibodies which can provide a momentary view (snapshot) of cTn levels but the current modality highly restricts access to and frequency of testing due to the requirement of a blood draw [34]. Frequent blood sampling may be uncomfortable and discourage ongoing monitoring, complicating immediate clinical decision-making [35]. Therefore, the development of novel techniques and patient-friendly approaches with the exploration of alternative mediums, which have the potential to enable more accessible, frequent and non-invasive detection of myocardial injury and its accurate assessment in real-time, including under exercise conditions, is imperative.

With the recent release of the saliva proteome, testing of oral fluid samples has become increasingly popular for the diagnosis of a variety of oral and systemic diseases [36–39]. Indeed, saliva as a non-invasive diagnostic medium is well-suited to POC applications due to its ease of collection, storage, and overall fluid management [13, 35, 39]. Recent studies have revealed that cardiac-specific troponin molecules are able to pass (filter) through the blood-salivary barrier into saliva [40, 41]. Furthermore, as evidence of the existence of the transport mechanism for the removal of cardiac-specific troponins across the blood-salivary barrier, few clinical trials have reported on the correlation between salivary and serum concentrations of cTn in patients with AMI and ischemic heart disease [40–44]. However, the findings have not yet been translated into actual clinical practice. One inherent challenge associated with saliva is that cTn concentrations in saliva are typically lower than those in serum [13]. Thus, more detailed, large-sample and statistically based studies are needed to determine a clinically meaningful threshold for defining elevated cTn in saliva. While both cardiac troponin-I (cTnI) and cardiac troponin-T (cTnT) have been accepted in various studies for their utility in identifying cardiac injury [19–29, 31–34], assessment of cTnI in saliva may be preferable because of its molecular mass which is almost 1.5 times less than that of cTnT due to distinct amino acid compositions and structural characteristics. As a result, cTnI is able to pass through the blood-salivary barrier more readily [40, 41].

It is reported here for the first time the measurements of post-exercise cTnI concentrations in saliva, exploring the possibility of non-invasive detection of exercise-induced myocardial injury. The purpose of this study was to compare cTnI concentrations in saliva and serum in male runners before, early, 4 h, and 24 h after the 5-km time-trial. Athletes who did not undergo exercise testing were recruited as controls for the present study. Choice of this control group was expected to amplify potential differences between salivary samples of exercising and non-exercising individuals and thus allow for more efficient identification of exercise-induced changes in cTnI levels in saliva for use in subsequent larger studies.

## Methods

### Participants and study design

A prospective, observational cohort study, in which cTnI levels in saliva and serum are compared, was implemented at Lobachevsky University. 82 well-trained male runners (age: 26.82±2.49 years; height: 174.26±2.71 cm; body mass: 67.94±2.64 kg; body mass index: 22.38±0.99 kg/m^2^) with at least three years’ regional-level competition experience were recruited. All participants were systemically healthy, asymptomatic, normotensive, non-smokers, and free of medication. Based on the pre-participation screening evaluations, they had no significant medical history and were judged to be free of cardiovascular disease (or other limiting conditions). Exclusion criteria for all volunteers were respiratory infection, use of steroidal medications, orthopedic injury (sprain, bruise or fracture), inability to provide a blood or saliva sample, fair or poor oral health, and age <18 years.

The study was approved by the Bioethics Committee of Lobachevsky University (Nr. 43) and was conducted in accordance with the guidelines laid down in the Declaration of Helsinki [45]. All participants gave written informed consent prior to enrollment in this study, after thorough explanation of the study design and protocol. After signing informed consent, they experienced medical screening in the laboratories of Lobachevsky University to meet eligibility criteria. Oral health was assessed by a registered dental hygienist in the dental operatory. Oral health was scored as poor, fair, or good based on the presence or absence of dental complaints, degree of mucosal inflammation, and extent of decay and periodontal diseases, which were determined by a single examiner per a modification of the oral health scoring system [46]. All participants were instructed to have breakfast 1 h before exercise testing and to refrain from ingesting caffeine and alcohol for at least 24 h prior to saliva and blood collection.

All participants comprised long-distance runners who kept a typical training routine. Weekly training was at least ≥3 sessions, for at least ≥2 h per training session. Exercise testing, which consisted of a 5-km time-trial, was conducted in the morning, and all runners did not exercise for 24 h before it. To determine whether exercise can affect salivary and serum levels of cTnI, all participants were divided into two groups. Of 82 consecutive runners recruited, 54 participants (group 1) completed a 5-km time-trial at top speed around a 400 m athletics track, while 28 participants (group 2, non-competitive controls) did not undergo this exercise testing and sat on chairs during it at the Athletics Centre. Runners in group 1 were encouraged to produce maximal effort, and their times were manually recorded using a stopwatch. Participants of both groups were examined for saliva and blood sampling at four time points (pre-exercise, post-exercise, 4 h post-exercise, and 24 h post-exercise), followed by a measurement of cTnI levels.

### Blood and saliva sampling

Blood (4 mL) was drawn from the median basilic vein and put into vacutainers containing anticoagulant. Unstimulated saliva was collected into sterile tubes for five min using the spitting method. Prior to saliva sampling, all participants thoroughly rinsed their mouths and swallowed the remaining water. Each specimen of blood and saliva was obtained at four time points as follows: pre-exercise (no more than fifteen min before exercise testing), post-exercise (within fifteen min after the 5-km time-trial), 4 h post-exercise, and 24 h post-exercise. All samples were transported on ice to a dedicated core laboratory at Lobachevsky University, centrifuged at 3000 rpm for fifteen min, divided into aliquots, and frozen at −80 °C until the time of cTnI measurement. Analyses were performed using the POC testing device (Getein1100, Getein Biotech Inc., China) in a blinded fashion in the laboratory within 3 months of storage.

### Device description

The POC-cTnI-Getein1100 assay (Cardiac Troponin I Fast Test) is a quantitative immunochromatographic assay which uses an anti-human cTnI monoclonal antibody conjugated with fluorescence latex and another anti-human cTnI monoclonal antibody coated on the test line. Serum or saliva samples were applied to the sample well. After the sample has been applied to the test strip, the fluorescence latex-labelled anti-human cTnI monoclonal antibody binds with the cTnI in a sample and forms a marked antigen-antibody complex. This complex moves to the test card detection zone by capillary action. Then marked antigen-antibody complex is captured on the test line by the anti-human cTnI monoclonal antibody. The fluorescence intensity of the test line increases in proportion to the amount of cTnI in a sample. The on-instrument turnaround time for the POC-cTnI-Getein1100 assay is 10 min. The measuring range for the POC-cTnI-Getein1100 assay is 0.10∼50.00 ng/mL. The lower detection limit, within-run precision, and between-run precision are as follows: ≤0.10 ng/mL, ≤10%, ≤15%, respectively, according to the manufacturer. The POC-cTnI-Getein1100 assay was previously compared with the Siemens ADVIA cTnI assay and the correlation coefficient (r) was 0.99.

### Statistical analysis

Data in the text are reported as mean ± standard deviation (SD). Data in Figure 1 are expressed as a median and interquartile range. Data in Figure 2 are expressed as mean ± SD, augmented with the median and interquartile range. The assumption of normality was estimated by using a Shapiro–Wilk test. Given that all data were normally distributed, between-group differences were examined using Student’s t-test, augmented with the calculation of respective effect sizes. Considering the sample size and given a power of 80% with an α of 0.05, an effect size of Cohen’s d = 0.77 would be needed to determine between-group differences in cTnI levels in both saliva and serum. Because there was no homogeneity of the variances, as evidenced by the results of the Fligner–Killeen test, within-group differences were examined using the Friedman test, with a Durbin–Conover pairwise post-hoc test in case of significant differences. Pearson’s correlation coefficient was used to indicate a significant linear relationship between salivary and serum levels of cTnI, and the ordinary least squares (OLS) regression line, Deming regression line, and Passing–Bablok regression line were displayed in the scatter plots. The Bland–Altman method of differences was applied to determine both a differential bias (i.e., a constant difference between salivary and serum levels of cTnI) and a proportional bias (i.e., a difference that depends on the value of the mean of salivary and serum levels of cTnI). Since the differences between salivary and serum levels of cTnI were normally distributed, the assumptions of the Bland–Altman method were met [47, 48]. To visually evaluate the performance of the new measurement approach (i.e., measurement of cTnI levels in saliva) against the reference method (i.e., measurement of cTnI levels in serum), the MethodCompare R package, which produces the “bias plot”, the “precision plot”, and the “comparison plot”, was used [49]. The MethodCompare R package implements a new statistical methodology, first introduced by Taffé [50], to identify and quantify the amounts of differential and proportional biases in the setting of heteroscedastic measurement errors, particularly when heteroscedasticity is a function of the latent trait. According to the MethodCompare R package requirements, six replicates were made with the reference standard for each of 10 randomly selected individuals in group 1, and five replicates with the new measurement approach. A value of *p* < 0.05 was considered as significant. The Holm correction to adjust the *p* value was applied, if required. All statistical tests were performed using RStudio software, version 2022.07.2+576 for macOS (RStudio, PBC, Boston, MA; http://www.rstudio.com).

**Figure 1.**
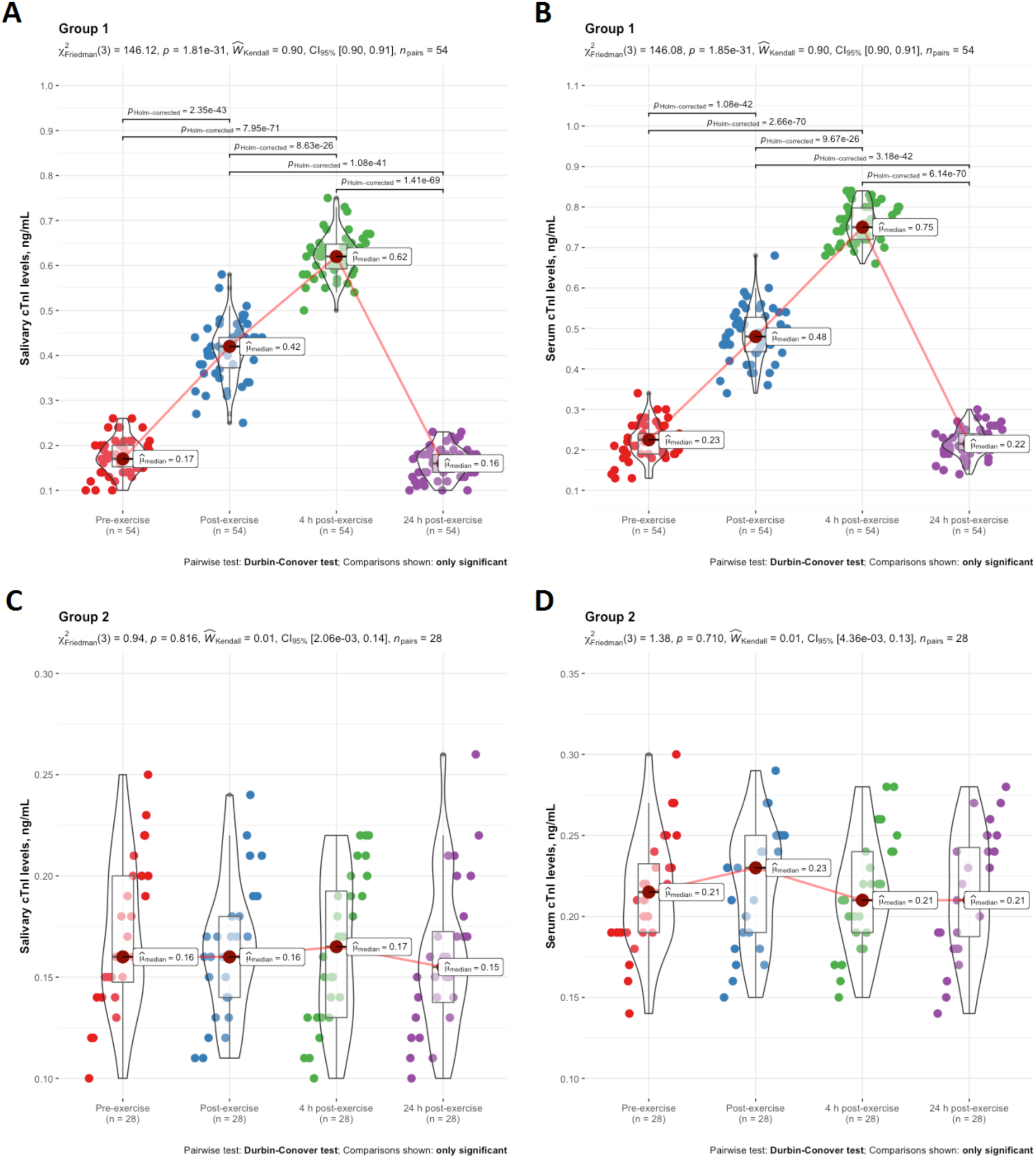
Salivary and serum levels of cTnI in participants of group 1 (A, B) and group 2 (C, D) at pre-exercise, post-exercise, 4 h post-exercise, and 24 h post-exercise. Data are expressed as a median and interquartile range, and are compared using the Friedman test, with a Durbin– Conover pairwise post-hoc test in case of significant differences. cTnI, cardiac troponin-I.

**Figure 2.**
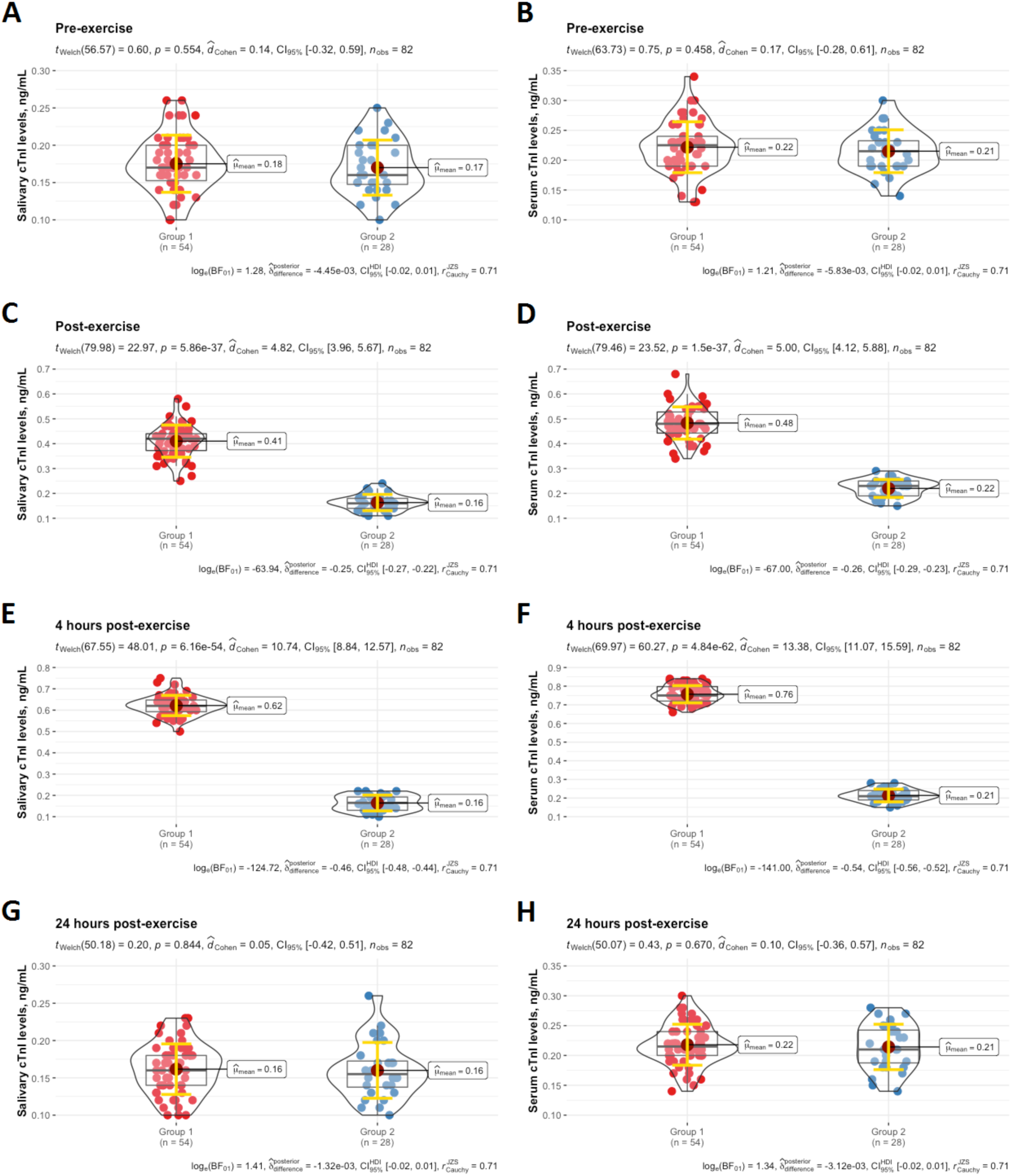
Salivary and serum levels of cTnI in participants of both groups at pre-exercise (A, B), post-exercise (C, D), 4 h post-exercise (E, F), and 24 h post-exercise (G, H). Data are expressed as mean ± SD augmented with the median and interquartile range, and are compared by Student’s t-test for independent data for between-group comparison analysis. cTnI, cardiac troponin-I.

## Results

All participants tolerated the study well and no adverse events were reported during and following the 5-km time-trial in group 1. Times to complete the 5 km for the runners in group 1 were 1089.85 ± 43.87 s.

Salivary and serum concentrations of cTnI were longitudinally evaluated across four time points in both groups. In group 1, as shown in Figure 1, salivary and serum levels of cTnI increased immediately after exercise compared to pre-exercise, reaching the highest values 4 h post-exercise with the subsequent return to baseline values 24 h post-exercise. In participants of group 2 who sat on chairs at the Athletics Centre during the 5-km time-trial, there were no time-dependent changes in cTnI levels in both saliva and blood serum up to 24 h after exercise. Salivary and serum concentrations of cTnI were significantly lower in group 2 compared to group 1 immediately after exercise and 4 h post-exercise (Figure 2). There was no difference in salivary and serum levels of cTnI between groups at pre-exercise and 24 h post-exercise.

Significant positive correlation was observed between cTnI levels in saliva and serum in both groups at all four time points (Figure 3). The highest correlation coefficient was verified between salivary and serum levels of cTnI in group 1 at 4 h post-exercise. Deming regression and Passing–Bablok regression revealed that there was a proportional agreement (i.e., the 95% confidence interval (CI) for the slope contained the value 1) between salivary and serum levels of cTnI in both groups at all four time points, whereas there was a significant differential bias (i.e., the 95% CI for the intercept did not contain the value 0) between the measurements in saliva and serum in both groups at least 4 h post-exercise.

**Figure 3.**
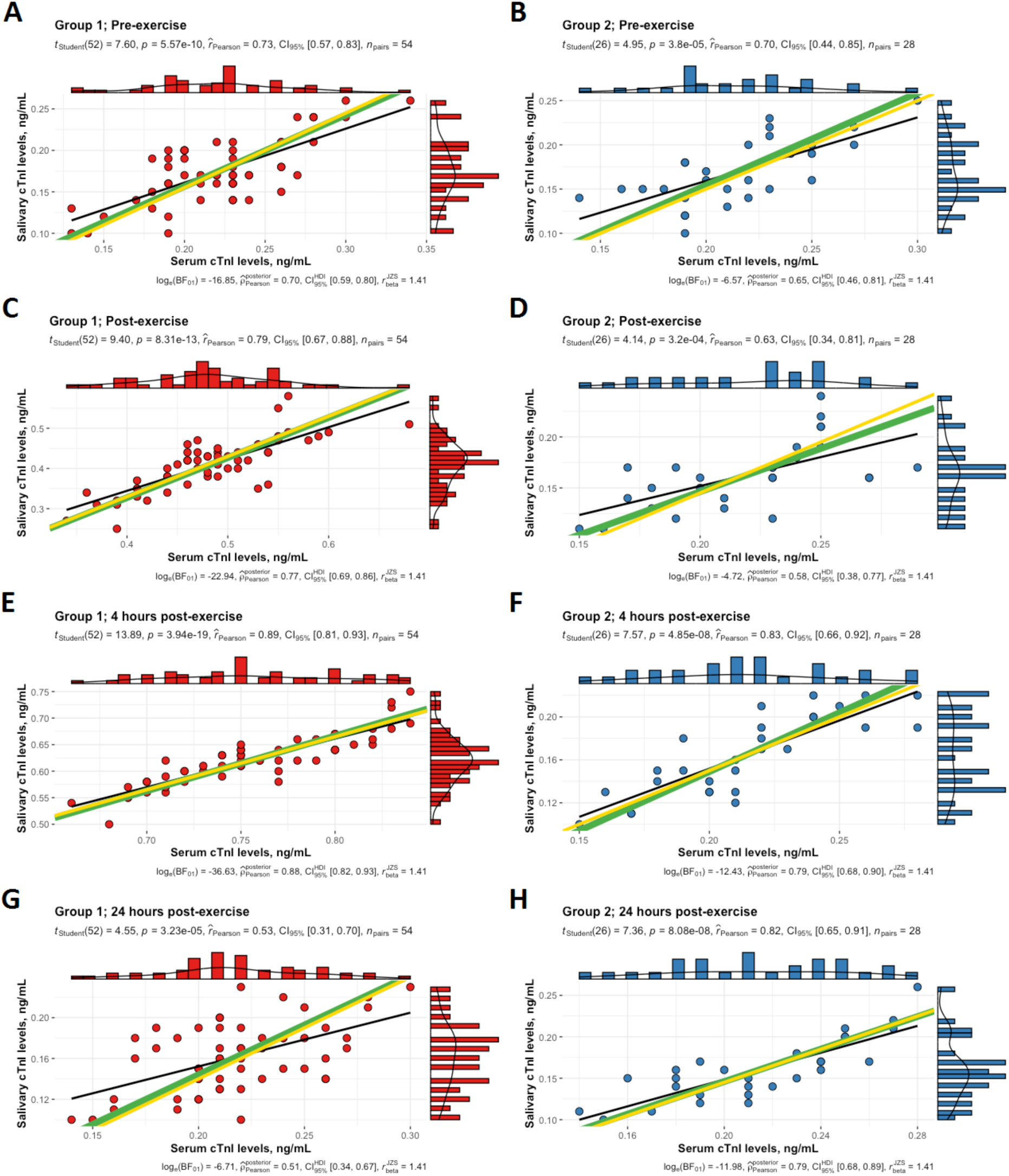
Correlation between salivary and serum levels of cTnI in participants of group 1 (A, C, E, G) and group 2 (B, D, F, H) at pre-exercise, post-exercise, 4 h post-exercise, and 24 h post-exercise. The solid black lines represent the ordinary least squares regression of salivary cTnI levels on serum cTnI levels. The solid green and yellow lines represent the Deming regression and Passing–Bablok regression, respectively. cTnI, cardiac troponin-I.

The Bland–Altman method of differences indicated that there was a negative differential bias in the data because the line of equality was higher than the 95% CI of the mean difference between salivary and serum levels of cTnI in participants of both groups at all four time points (Figure 4). However, there was no proportional bias in the Bland–Altman plots, since the slope of the OLS regression of differences on means did not differ significantly from the value 0 (p > 0.05) and, equivalently, the 95% CI for the slope contained the value 0. Therefore, the limits of agreement were constructed without first finding a suitable transformation of the data. All the data points, except two points on the Bland–Altman plot created using the values of cTnI in group 1 at post-exercise (Figure 4C), were observed inside ±1.96 SD of the mean difference. In Figure 5, the extended Bland–Altman limits of agreement plot for each time point indicates that there was a negative differential bias of the new measurement approach (i.e., measurement of cTnI levels in saliva) for the values of the estimated latent trait level (i.e., the best linear unbiased prediction (BLUP) of *x*). The bias plots illustrated that the bias was negative for all the values of the estimated latent trait level (i.e., BLUP of *x*). In addition, it can be seen on the bias plots created for all four time points that the bias was more negative for higher values than for lower values of the true latent trait. The precision plots showed that after recalibration the new measurement approach (i.e., measurement of cTnI levels in saliva) was less precise than the reference standard (i.e., measurement of cTnI levels in serum) in general. However, the new measurement approach (i.e., measurement of cTnI levels in saliva) had similar (almost equal) precision to the reference method (i.e., measurement of cTnI levels in serum) at the following time points: post-exercise, 4 h post-exercise, 24 h post-exercise. Of interest is to note that the variance of the measurement errors of both measurement approaches (i.e., measurement of cTnI levels in saliva and serum) was almost constant throughout the whole range of the true latent trait. One can see on the comparison plots created for all four time points that recalibration of the new measurement method (i.e., measurement of cTnI levels in saliva) was effective in removing existing bias.

**Figure 4.**
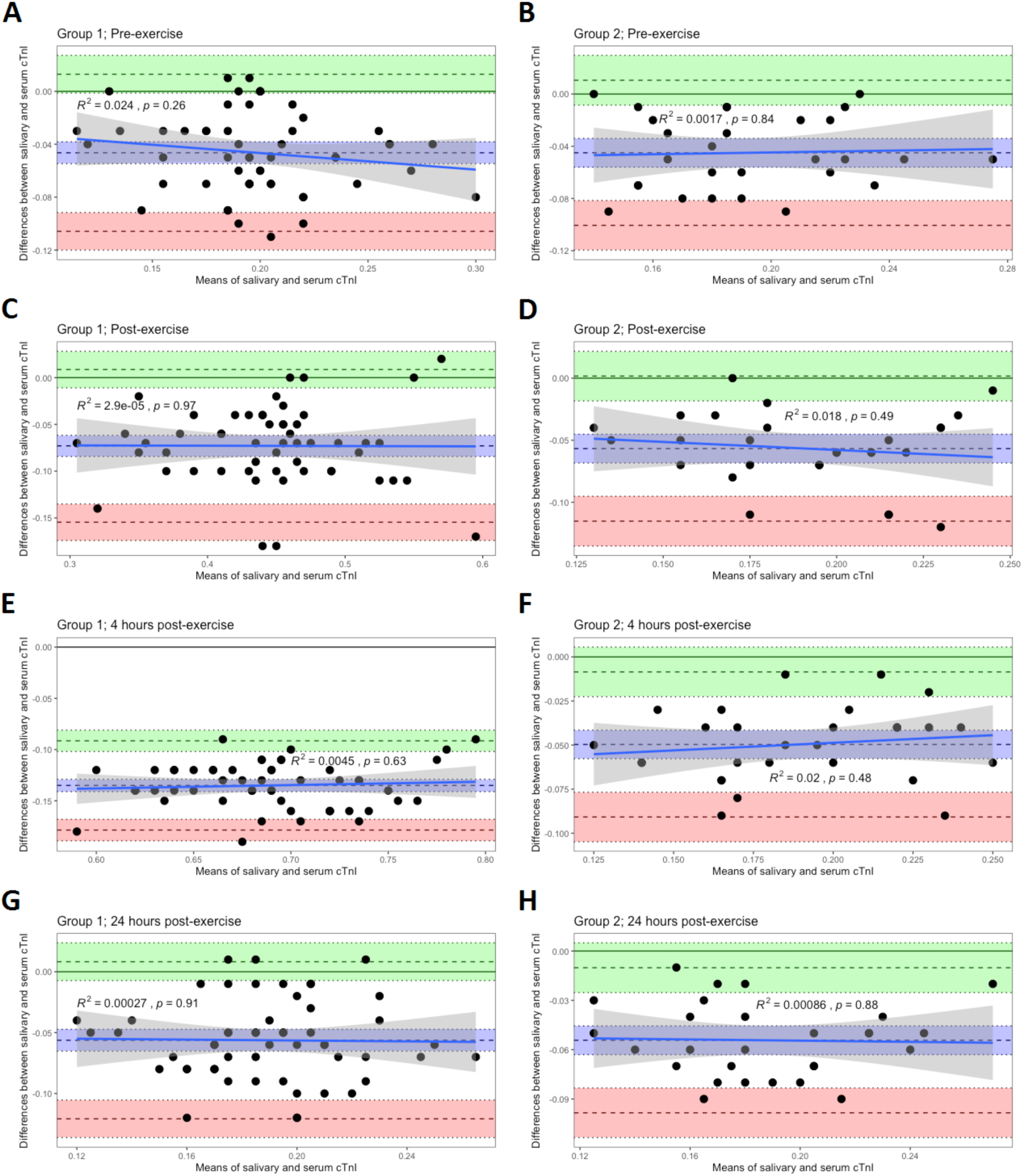
Bland–Altman plots of differences against means for comparisons of salivary and serum levels of cTnI in participants of group 1 (A, C, E, G) and group 2 (B, D, F, H) at pre-exercise, post-exercise, 4 h post-exercise, and 24 h post-exercise. The mean difference is represented by a dashed line (inside the blue area) parallel to the *x* axis. The limits of agreement are represented by dashed lines parallel to the *x* axis at −1.96 SD (inside the red area) and +1.96 SD (inside the green area). Shaded areas represent the 95% confidence interval limits for the mean difference (blue shading) and for the agreement limits (red and green shading). The solid blue lines represent the ordinary least squares regression of differences on means. The grey shaded regions indicate 95% confidence intervals. cTnI, cardiac troponin-I.

**Figure 5.**
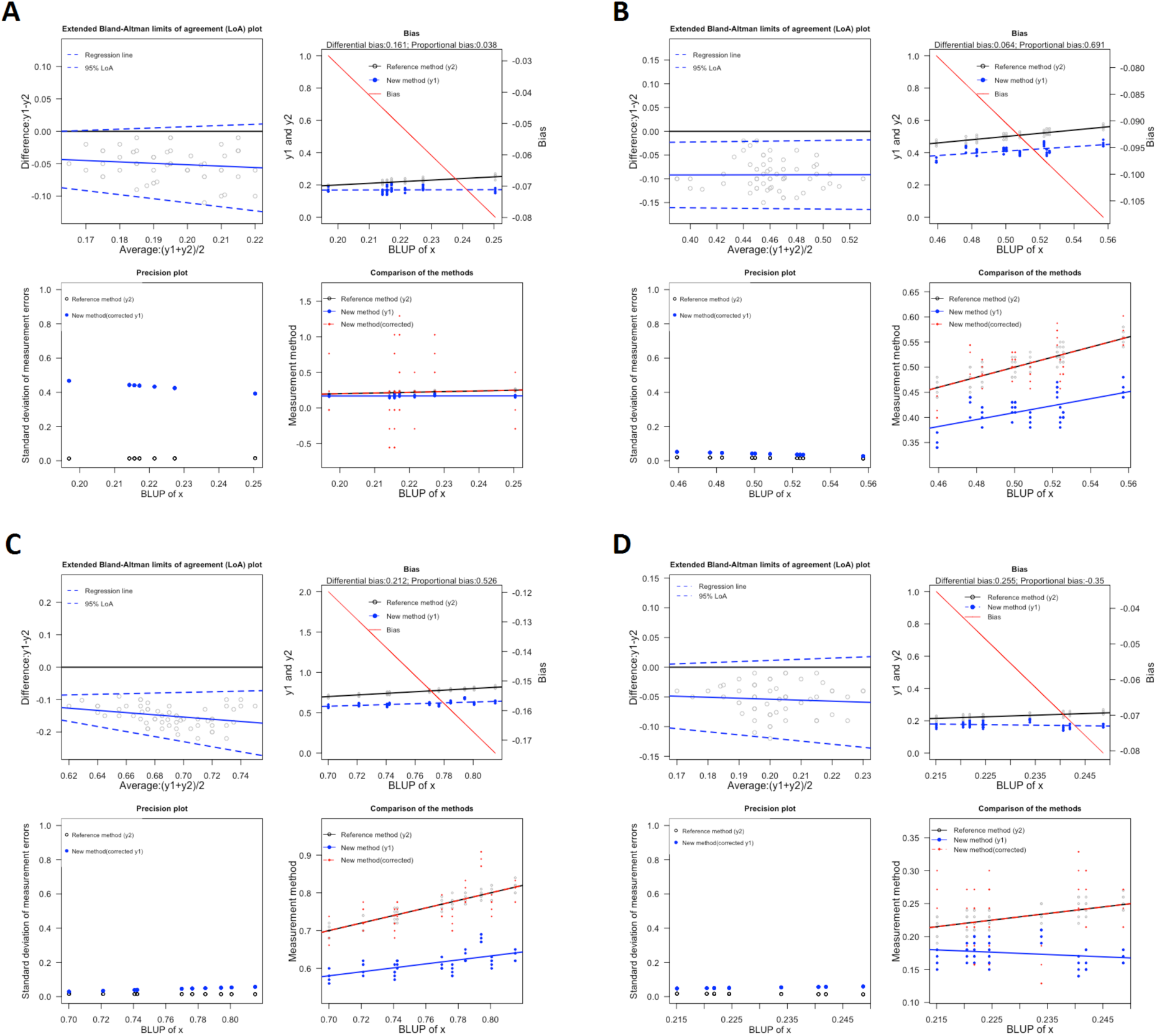
Generalized plots created using the MethodCompare R package for the following time points: pre-exercise (A), post-exercise (B), 4 h post-exercise (C), 24 h post-exercise (D). Each generalized plot contains (top left) the extended Bland–Altman limits of agreement plot, (top right) bias plot showing the amount of bias of the new measurement method, (bottom left) precision plot showing the precision (i.e., standard deviation of the measurement error) of each measurement method, and (bottom right) scatter plot illustrating that the recalibration of the new measurement method (i.e., corrected) was effective.

Within-run precision of the POC-cTnI-Getein1100 assay for the saliva and serum samples of 10 randomly selected individuals in group 1 was less than 10%, 5%, 3%, and 10% at pre-exercise, post-exercise, 4 h post-exercise, and 24 h post-exercise, respectively, as evidenced by assaying at least 5 replicates.

## Discussion

This is the first study to compare exercise-induced changes in salivary and serum levels of cTnI in male athletes. Pattern of cTnI alterations caused by the 5-km time-trial was similar in saliva and blood serum, as demonstrated by quantifications of cTnI across multiple time points. Although there was a proportional agreement between salivary and serum levels of cTnI in both groups at all four time points, there was also a negative differential bias of the new measurement approach (i.e., measurement of cTnI levels in saliva) compared to the reference method (i.e., measurement of cTnI levels in serum). In other words, cTnI levels in saliva were constantly lower than those in serum in both groups at all four time points. However, recalibration of the new measurement method (i.e., measurement of cTnI levels in saliva) by using the MethodCompare R package [49] was effective in removing existing bias, as evidenced by its similar precision to the reference method (i.e., measurement of cTnI levels in serum), particularly at the following time points: post-exercise, 4 h post-exercise, and 24 h post-exercise.

The exact mechanisms behind the movement of cTnI between blood serum and saliva are not yet fully understood. Recent clinical trials utilizing troponin immunoassays have provided clear evidence that cardiac-specific troponin molecules are capable of crossing the glomerular filtration barrier [41, 51]. This finding suggests that transfer of cardiac-specific troponins into oral fluid via the blood-salivary barrier may be mediated by a parallel transport mechanism. Given that cTnI was previously identified in saliva, in which its levels were permanently lower than those in blood serum, passive diffusion to the oral fluid through the blood-salivary barrier may be one of the mechanisms for the removal of cardiac-specific troponin-I and its fragments from the bloodstream [40–41]. Supporting this notion, the correlation between salivary and serum levels of cTnI in AMI patients has been demonstrated in a number of pilot studies [40–44]. Furthermore, exercise-induced oxidative damage to lipids in the plasma membrane of salivary gland cells and/or blood vessel cells could change membrane permeability [52–54], prompting passive transport of cTnI and its fragments from the circulation to the oral fluid through the blood-salivary barrier.

Release kinetics of cTnI elevations following exercise are less clear than those after AMI [20]. Middleton et al. reported a pattern of biphasic cTnT release into the circulation during and after a marathon run [55]. Because cTnI levels in both serum and saliva were not quantified during exercise in the present study, the potential for a biphasic release of cTnI with its corresponding elevated state in saliva both during and after the 5-km time-trial cannot be supported or rejected. However, the data of this study are the first to demonstrate that salivary concentrations of cTnI progressively increased in all participants in group 1 after exercise cessation with peak values occurring 4 h after completion of a 5-km time-trial. The magnitude of exercise-induced cTnI elevations in both saliva and serum varied among participants in group 1 with some individuals demonstrating small changes in cTnI concentrations, but others reporting values exceeding several times the upper reference limit. Importantly, exercise-induced increases in salivary and serum levels of cTnI were transient in all participants in group 1, with values returning to baseline after 24 h post-exercise. Although there are no available data to confirm the results of this study describing the kinetics of exercise-induced cTnI concentrations in saliva, the observations regarding post-exercise cTnI concentrations in serum in the present study are consistent with findings from several other studies that assessed time-dependent changes in serum levels of cTnI up to 48 h after various types of exercise [56–59].

The mechanisms responsible for post-exercise cTnI elevations remain controversial [20]. The European Society of Cardiology’s Study Group on Biomarkers identified three possible causes for elevated cTn concentrations [60]: (i) reversible damage attributable to cell wounds, increased exocytosis rates, and release of extracellular blebs; (ii) cardiac damage attributable to apoptosis; and (iii) irreversible damage attributable to myocardial necrosis. While direct data to support or refute a single release mechanism for exercise-induced increases in cTn concentrations are limited, the following explanations based on available evidence can nonetheless be made regarding the underlying mechanisms. First, stressing the cardiomyocytes via contraction, β-adrenergic stimulation, stretching, and/or brief ischemia during exercise may alter their sarcolemmal permeability [61–64], leading to passive diffusion of cTnI and its fragments from the cardiomyocyte to the extracellular space and circulation [65]. Smaller fragments of cTnI may leak directly into the bloodstream, while its larger fragments might only escape with modification or destruction of the cell membrane [20]. Second, acute bouts of exercise may increase apoptotic rates, whereas increased left ventricular preload and/or brief ischemia (conditions that can occur during exercise) may also induce isolated apoptosis [66–70]. On the one hand, apoptosis should not cause cTnI elevations outside the cardiomyocyte because intracellular content is not released when the apoptotic cell is fragmented and engulfed by other cells, but cTnI could be released by destroying apoptotic bodies or transitioning from cardiomyocyte apoptosis to secondary necrosis [71]. Third, an acute bout of exercise may accelerate cardiomyocyte turnover, which might lead to cTnI release into the bloodstream from replaced cardiomyocytes [20, 72, 73]. Finally, given that the cTnI release following exercise is smaller, peaks earlier, and resolves more rapidly than after AMI, necrosis is unlikely to be responsible for exercise-induced cTnI increases, but these differences do not exclude the possibility that a small degree of necrosis may occur and contribute to increased cTnI levels in vulnerable individuals after exercise [20]. Nonetheless, there is no evidence of incident myocardial edema or late gadolinium enhancement in runners directly following exercise [74, 75]. However, imaging techniques may lack enough sensitivity to detect a very small degree of necrosis with associated elevations in cTnI levels in the circulation [76].

A number of alternative hypotheses, which have been proposed to explain exercise-induced increases in cTnI levels, may also be taken into consideration with some limitations. First, it is suggested that exercise-induced dehydration may affect post-exercise cTnI levels in serum and saliva, but the percentage change in fluid balance markers is usually (very) small in relation to elevations in cTnI concentrations [20]. In addition, any dehydration is expected to be restored soon by post-exercise rehydration, which is contradictory to the progressive elevations in cTnI levels in serum and saliva up to 4 h after exercise. Moreover, hemodilution may occur after endurance exercise [77], so the impact of hemoconcentration on post-exercise cTnI levels in blood serum is likely limited if not negligible. Second, evidence suggests that exercise-related mild reduction in kidney function may attenuate renal elimination of cTnI, leading to an increase in its levels in the circulation following exercise [78]. However, cTnI elevations caused by prolonged exercise typically far exceed the modest attenuation in kidney function, indicating that the contribution of exercise-induced reduction in renal function to the magnitude of post-exercise cTnI concentrations in the bloodstream is limited [20]. Third, few studies reported evidence of cross-reactivity of cTn assays with skeletal troponin or muscle injury with cTn release, but this is limited to cTnT assays in patients with rhabdomyolysis and skeletal myopathies and not to athletes or cTnI assays [79–82]. Given that cTnT, unlike its cTnI counterpart, is detected in both diseased skeletal muscle and myocardium, muscle damage may presumably contribute to exercise-induced cTnT elevations but probably might not contribute to the magnitude of cTnI increases [83].

Taken together, it is logical to assume that exercise-induced cTnI elevations observed in this study are mainly attributable to reversible membrane damage to viable cardiomyocytes. Whether this was the only mechanism responsible for exercise-induced cTnI increases in both serum and saliva, or there was also isolated apoptosis and/or a very small degree of necrosis, is unknown in the present study. Moreover, it is unclear whether the supposed alterations in membrane permeability are fully physiological (part of a remodeling process). Given that the magnitude of cTnI elevations was variable across participants in group 1 after the same exercise it is possible that the contribution of these underlying mechanisms to the observed variability differed between athletes with various post-exercise levels of cTnI.

In a sports and exercise medicine setting, quantification of cTnI levels in saliva utilizing the POC-cTnI-Getein1100 assay may be a useful non-invasive tool in evaluating whether exercise-induced increases in cTnI levels are transient or there are acutely or chronically elevated cTnI concentrations. Consequently, the proposed non-invasive POC assay of cTnI in saliva may be envisioned to empower practicing cardiologists and sport physicians in determining timely measures to prevent unnecessary exercise-induced myocardial injury in real time. However, despite the early peak and relatively low absolute values, persistent increases in cTnI concentrations up to 4 h after exercise mean that 4-h sequenced saliva collections likely cannot be used to differentiate exercise-induced cTnI release from early-onset cardiac events. Therefore, the pattern and magnitude of exercise-induced cTnI elevations in saliva should be interpreted with caution, especially during or immediately after acute bouts of endurance exercise when suspected cardiac events may occur.

### Study limitations

First, our findings cannot easily be generalized to all athletes participating in various sports because only well-trained male runners were enrolled in this study. Second, although all laboratory procedures were performed according to stringent standardized operating procedures, human error in the handling of study-specific saliva/blood samples may have occurred in a small number of samples leading to incorrect results pertaining to an individual athlete. This would invariably have led to an underestimation or overestimation of the true precision of the new measurement approach (i.e., measurement of cTnI levels in saliva).

### Conclusions

This is the first study to report a proportional agreement between salivary and serum concentrations of cTnI in male athletes before, early, 4 h, and 24 h after the 5-km time-trial. Recalibration of the new measurement approach (i.e., measurement of cTnI levels in saliva) by using the MethodCompare R package was effective in removing existing bias, as evidenced by its similar precision to the reference method (i.e., measurement of cTnI levels in serum). Consequently, saliva as a diagnostic tool offers exciting potential in athletic settings by providing credible assessment of exercise-induced cardiac damage without the need for collecting blood. Furthermore, the proposed assay of cTnI in saliva opens novel possibilities for the development of a biosensor to monitor time-dependent changes in post-exercise cTnI concentrations in a non-intrusive manner.

### Perspectives

#### Competency in systems-based practice

After recalibration, the POC-cTnI-Getein1100 assay utilizing saliva has precision comparable to that of an assay using serum and can be used to non-invasively identify post-exercise cTnI release and exercise-induced myocardial injury in male runners.

#### Translational Outlook

Larger studies and prospective trials are needed to compare the value of implementing the POC-cTnI-Getein1100 assay utilizing saliva to that of conventional strategies in the assessment of exercise-induced myocardial injury in athletes participating in different sports, as well as vulnerable individuals.

## Funding

This study was funded by the Russian Science Foundation [project number 23-75-01039, https://rscf.ru/en/project/23-75-01039/].

## Declaration of competing interest

The author declares that he has no known competing financial interests or personal relationships that could have appeared to influence the work reported in this paper.

## Acknowledgments

The author appreciates male runners and their coaches for their collaboration and commitment to this study.

## CRediT authorship contribution statement

**Aleksandr N. Ovchinnikov:** Conceptualization, Data curation, Formal analysis, Funding acquisition, Investigation, Methodology, Resources, Supervision, Visualization, Writing – original draft, Writing – review & editing.

## Data availability statement

Data will be made available on request.

## Abbreviations and Acronyms

AMI: acute myocardial infarction
CI: confidence interval
cTn: cardiac troponin
cTnI: cardiac troponin-I
cTnT: cardiac troponin-T
OLS: ordinary least squares
POC: point-of-care
SD: standard deviation
SCD: sudden cardiac death

## Notes

### Competing Interest Statement

The authors have declared no competing interest.

### Author Declarations

Bioethics Committee of Lobachevsky University gave ethical approval for this work.

